# Detection of SARS-CoV-2 antibodies formed in response to the BNT162b2 and mRNA-1237 mRNA vaccine by commercial antibody tests

**DOI:** 10.1101/2021.03.30.21254604

**Authors:** Jamil N Kanji, Ashley Bailey, Jayne Fenton, Sean H Ling, Rafael Rivera, Sabrina Plitt, Wendy I Sligl, Sean Taylor, LeeAnn Turnbull, Graham Tipples, Carmen L. Charlton

## Abstract

**PURPOSE:** With rapid approval of SARS-CoV-2 vaccines, the ability of clinical laboratories to detect vaccine-induced antibodies with available high-throughput commercial assays is unknown. We aimed to determine if commercial serology assays can detect vaccine-induced antibodies (VIAs) and understand the vaccination response.

**METHODS:** This cohort study recruited healthcare workers and residents of long-term care facilities (receiving the BNT162b2 and mRNA-1273 products, respectively) who underwent serum collection pre-vaccination (BNT162b2 group), 2-weeks post vaccination (both groups), and pre-2^nd^ dose (both groups). Sera were tested for the presence of SARS-CoV-2 IgG using four commercial assays (Abbott Architect SARS-CoV-2 IgG, Abbott Architect SARS-CoV-2 IgG II Quant, DiaSorin Liaison Trimeric S IgG, and GenScript cPASS) to detect VIAs. Secondary outcomes included description of post-vaccination antibody response and correlation with neutralising titers.

**RESULTS:** 225 participants (177 receiving BNT162b2 and 48 receiving mRNA-1273) were included (median age 41 years,; 66-78% female). Nucleocapsid IgG was found in 4.1% and 21.9% of the BNT162b2 (baseline) and mRNA-1273 (2-weeks post first dose). All anti-spike assays detected antibodies post-vaccination, with an average increase of 87.2% (range 73.8-94.3%; BNT162b2), and 25.2% (range 23.8-26.7%; mRNA-1273) between the first and last sampling time points (all p<0.05). Neutralising antibodies were detected at all post-vaccine timepoints for both vaccine arms, with increasing titers over time (all p<0.05).

**CONCLUSION:** Anti-spike vaccine-induced SARS-CoV-2 IgG are detectable by commercially available high-throughput assays and increases over time. Prior to second dose of vaccination, neutralising antibodies are detectable in 73-89% of individuals, suggesting the majority of individuals would have some degree of protection from subsequent infection.

## Introduction

To date, the principal utility of SARS-CoV-2 IgG high throughput serology assays has been to conduct seroprevalence studies for large cohorts to aid public health decision-making [1, 2]. Until recently, all seroprevalence studies have examined detection of antibodies from natural infection. While most vaccine candidates produced detectable antibody levels on specific serological research assays in early phase trials [3-5], it is unclear how currently available high-throughput commercial assays will detect vaccine-induced antibody in the clinical laboratory. In the case of varicella, detection of vaccine-induced antibody is variable among commercial kits compared to detection of antibodies produced from natural infection, which are more robust [6].

SARS-CoV-2 neutralization antibodies have been shown to predict disease severity and survival [7]. There is also evidence of their protective capacity in preventing infection in animal models and humans [8-10]. These data corroborate evidence for neutralising antibody (nAb) protection in numerous viral infections including yellow fever, encephalitis, dengue, mumps, influenza and others [11]. To date, there is little data measuring the level of nAbs pre/post-vaccination for SARS-CoV-2 nor how results compare with vaccine efficacy and protection.

Currently approved SARS-CoV-2 vaccines in the United States and Canada (BNT162b2 and mRNA-1273 products) consist of lipid nanoparticles formulated nucleoside-modified messenger RNA (mRNA) [12, 13]. Both formulations induce T-cell activation and production of antibodies in response to *in vivo* production of SARS-CoV-2 spike (S) protein following translation of the synthetic mRNA in human cells [14]. Antibodies against the receptor binding domain (RBD) found in the S1 region of the spike gene [3, 15], anti-S1 protein IgG [14], as well as neutralising antibody [14, 16] have been detected in response to both vaccines.

We aimed to evaluate the ability of three commercial SARS-CoV-2 IgG assays and one functional nAb test to detect and quantify antibodies in two separate patient populations receiving their first doses of the BNT162b2 and mRNA-1273 SARS-CoV-2 mRNA vaccines. It was hypothesized that assays targeting non-spike proteins (e.g., nucleocapsid (N) protein) would screen positive only in individuals who previously recovered from natural SARS-CoV-2 infection. It was further postulated that there could be a difference between the IgG binding antibody total immune response versus the nAb response to vaccination.

## Methods

### Participant sample collection

Serum samples were collected prospectively from two separate patient groups undergoing COVID-19 vaccination. The first group consisted of healthcare workers (HCWs) who received the BNT162b2 vaccine series while the second group consisted of residents of long-term care facilities who received the mRNA-1273 vaccine series. Herein the groups will be referred to as the BNT162b2 and the mRNA-1273 groups, respectively.

Serum samples in the BNT162b2 group were planned to be drawn at the following approximate time points: (i) at baseline (defined as 24-72 hours prior to the first dose, or up to five days post the first dose of vaccine), (ii) 14 days post first dose of vaccine; and (iii) within 24 hours of the second dose of vaccine (either the day before, day of, or day prior). Those in the mRNA-1273 group were planned to have samples collected at approximately (i) 14 days and (ii) 21-28 days post first dose of vaccine (pre-2^nd^ dose). Due to the rapid roll out of vaccine in long-term care facilities, none of the participants in the mRNA-1273 group had baseline/pre-first dose samples collected.

### Vaccine distribution

Details regarding Alberta COVID-19 vaccine distribution have been outlined previously [17]. Briefly, given the need for storage at -70°C, the BNT162b2 vaccine was provided to healthcare workers (those working in areas of intensive care, emergency, care of COVID-19 positive patients, and those working in long-term care) at a centralized vaccine depot. The mRNA-1273 product was transported for administration to residents of continuing and long-term care facilities (given ability to store at -20°C) [17]. The two doses of the BNT162b2 and mRNA-1273 products were administered three and four weeks apart, respectively as per vaccine manufacturer recommendations.

Vaccine administration and allocation was directed as per planned vaccine roll-out by provincial government health authorities and not by the researchers. Inclusion criteria to participate this study comprised being 18 years or older, eligible for and planned COVID-19 vaccine series. Participants who had recovered from previous SARS-CoV-2 infection, who were on biologics or other immunosuppressive medications, or who were pregnant were included.

### SARS-CoV-2 IgG detection

Samples were stored at -20°C and underwent one freeze-thaw cycle prior to testing. Sera were tested for SARS-CoV-2 IgG using three high-throughput commercial assays: Abbott Architect SARS-CoV-2 IgG (IgG targeting the N-protein; qualitative result only) [18], Abbott Architect SARS-CoV-2 IgG II Quant (IgG targeting the RBD-region of the S-protein; quantitative result) [18], and DiaSorin Liaison (trimeric S-protein assay targeting the S1/S2 regions of the S-protein; quantitative result) [19]. In addition, we evaluated the presence of SARS-CoV-2 nAbs using the GenScript cPass Neutralisation antibody detection kit (Table S1) [20, 21]. All testing was done according to product inserts and qualitative and quantitative values (where available) were recorded for each assay. Only the assay by GenScript has a claim to detect nAbs.

### Data analysis

Baseline demographics of all participants were extracted from the provincial electronic medical record (EMR) (Table S1). Continuous variables were compared using Mann-Whitney tests while categorical variables were compared using Chi-square or Fisher’s exact tests. All statistical analysis and graphing were done using Stata version 16.1 (Table S1).

## Results

### Population demographics

Serum from 225 participants who received an mRNA COVID-19 vaccine (177 receiving the BNT162b2 and 48 receiving mRNA-1273) (Table 1) was collected at up to three timepoints. The median age of all the participants was 41 years (with the mRNA-1273 having a significantly higher median age; 84 vs 37 years). The majority of participants in each group were female (66.7-78.1%). The median time between receipt of vaccine dose 1 and dose 2 for both groups was 25-26 days. A significantly higher proportion of mRNA-1273 participants had recovered from confirmed SARS-CoV-2 infection (33.3 vs 3.4%). Two participants in the BNT162b2 group were diagnosed with COVID-19 after receipt of their vaccine (one diagnosed seven days after the first dose; the other nine days after the second dose). Medical comorbidities were more commonly seen in the mRNA-1273 group. In total, five participants were being treated with biologics or immunosuppressive medications.

**Table 1.** Demographics by type of vaccine administered.

|  | Total | BNT162b2<br>group | mRNA-1273<br>group | p-value<sup>a</sup> |
| --- | --- | --- | --- | --- |
| N (%) | 225 (100) | 177 (78.7) | 48 (21.3) |  |
| Median Age (years; IQR) | 41 (31-55) | 37 (30-48) | 84 (75.5-90.5) | <0.001 |
| Gender (%) |  |  |  | 0.11 |
| Male | 55 (24.4) | 39 (22.0) | 16 (33.3) |  |
| Female | 170 (75.6) | 138 (78.0) | 32 (66.7) |  |
| Time between dose 1 and<br>dose 2 of vaccine (days) |  |  |  |  |
| Median (IQR) | 26 (25-27) | 26 (25-28) | 25 (24-26) | <0.001 |
| Recovered from COVID-19 | 22 (9.8) | 6 (3.4) | 16 (33.3) | <0.001 |
| Of these: |  |  |  |  |
| COVID-19 occurred prior<br>to first vaccine dose (%) | 20 (90.9) | 4 (66.7) | 16 (100) | 0.02 |
| Time from COVID-19<br>diagnosis to first vaccine<br>dose (median, range, days) | 45<br>(30.5-73) | 45.5<br>(36-82) | 43<br>(30.5-73) | 0.54 |
| COVID-19 occurred after<br>first vaccine dose (%) | 2 (0.9) | 2 (33.3) | 0 | - |
| Time from first dose to<br>COVID-19 diagnosis<br>(range, days) | 20.5 (7-34) | 20.5 (7-34) | 0 | - |
| Any Co-Morbidity or<br>Immunosuppression (%) | 45 (20.0) | 9 (5.1) | 36 (75.0) | <0.001 |
| Cardiac Disease (%) | 27 (12.0) | 0 | 27 (46.3) | <0.001 |
| Diabetes (%) | 12 (5.3) | 3 (1.7) | 9 (18.8) | <0.001 |
| Solid Cancer (%) | 5 (2.2) | 1 (0.56) | 4 (8.3) | 0.008 |
| Hematological Cancer (%) | 1 (0.44) | 1 (0.56) | 0 | 1.00 |
| Asthma (%) | 1 (0.44) | 1 (0.56) | 0 | 1.00 |
| Renal Disease (%) | 5 (2.2) | 0 | 5 (10.4) | <0.001 |
| COPD (%) | 8 (3.6) | 0 | 8 (16.7) | <0.001 |
| Dementia (%) | 22 (9.8) | 0 | 22 (45.8) | <0.001 |
| Transplant (%) | 0 | 0 | 0 | -- |
| Biologic or<br>immunosuppressant use<sup>b</sup><br>(%) | 5 (2.2) | 4 (2.3) | 1 (2.1) | 1.00 |
| Pregnant (%) | 0 | 0 | 0 | -- |

Abbreviations: COPD – chronic obstructive pulmonary disease; IQR – interquartile range
<sup>a</sup> p-value comparing BNT162b2 and mRNA-1273; calculated by Mann-Whitney test for age, time between vaccine doses and time from COVID-19 diagnosis to first vaccine variables, and chi-square or Fisher's exact test for remaining variables.
<sup>b</sup> Between both groups, five participants were considered immunosuppressed. In the BNT162b2 group, four participants were using biologics (adalimumab, daratumumab/bortezomib, dupilumab, infliximab) for ankylosing spondylitis (n=1), relapsed/refractory multiple myeloma (n=1), atopic dermatitis (n=1), and inflammatory bowel disease (n=1). One participant in the mRNA-1273 group used methotrexate for rheumatoid arthritis. Dupilumab is considered to have a low potential for immunosuppression.

### Anti-nucleocapsid IgG detection

IgG directed towards the SARS-CoV-2 N-protein was found in 3.6-5.4% and 18.9-21.9% of the BNT162b2 and mRNA-1273 groups, respectively (Fig. 1). Over the course of the blood sampling times in each group, the positivity rate for anti-N IgG detection did not change significantly from the baseline to pre-2^nd^ dose sample (*p*=0.86; BNT162b2 group) and from the 2-week and pre-2^nd^ dose samples (*p*=0.76; mRNA-1273 group) in either group.

**Figure 1.**
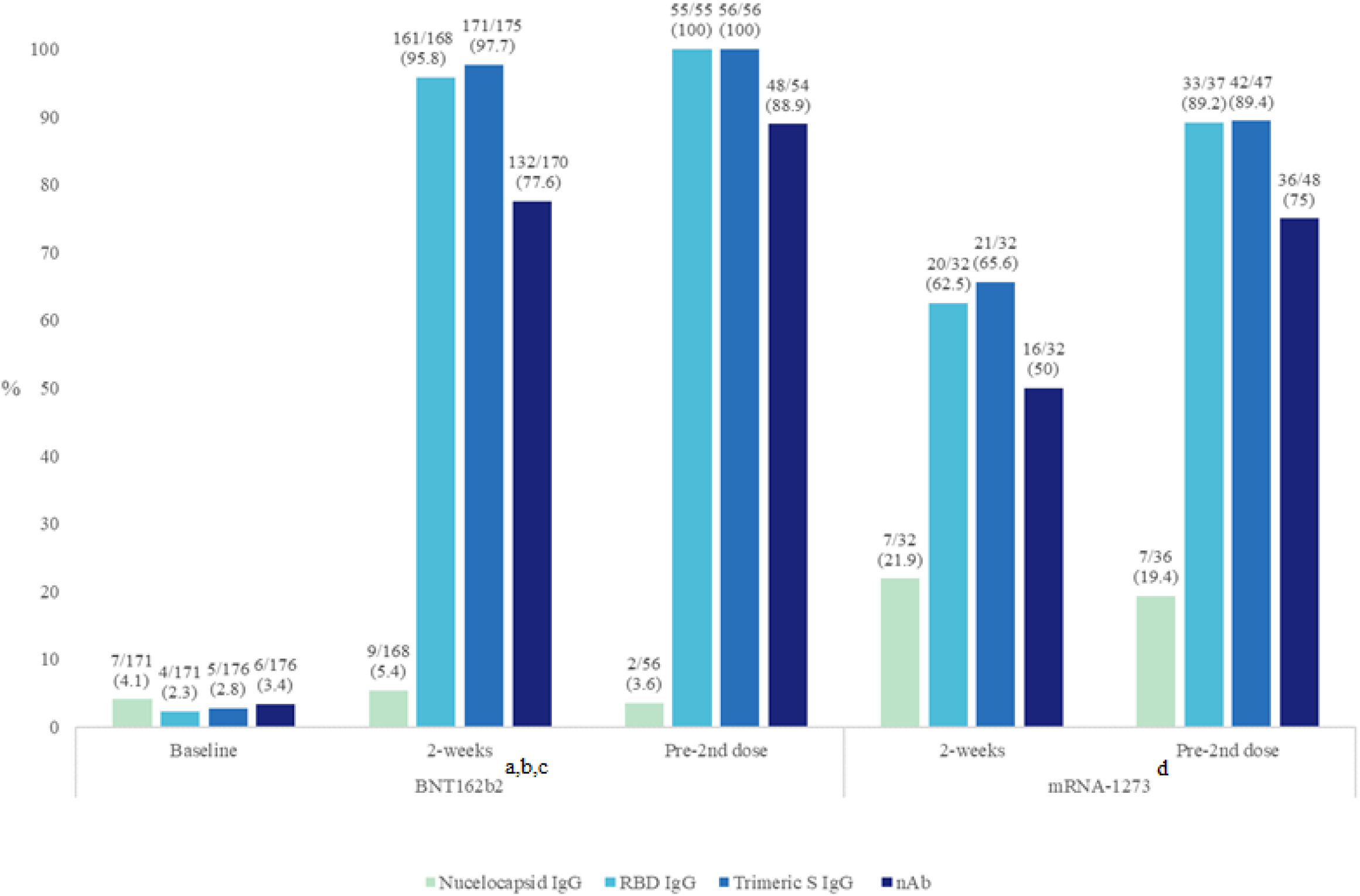
Positivity rates of each assay tested for each group at different time points. Positivity rates are presented in (%). ^a^Baseline versus 2 weeks comparison (BNT162b2): p=NS (Nucleocapsid IgG); p<0.0001 (RBD IgG, Trimeric S IgG, and nAb assays). ^b^2-week versus pre-2^nd^ dose comparison (BNT162b2): p=NS for all assays. ^c^All p-values comparing baseline to pre-2^nd^ dose (BNT162b2) vaccine were <0.0001, except for nucleocapsid IgG (p=1.0). ^d^2-weeks versus pre-2^nd^ dose comparison (mRNA-1273): p=NS (Nucleocapsid IgG); p<0.01 (RBD IgG); p<0.05 (Trimeric S IgG and nAb assays).

In the BNT162b2 group, all four participants who were COVID-19 recovered prior to their first dose had a detectable anti-N IgG (Table S2). One of four lost their anti-N IgG when tested two-weeks post first vaccine dose (60 days post diagnosis). 85.7% (6/7) COVID-19 recovered participants in the mRNA-1273 group had detectable anti-N IgG at 2-weeks post first vaccine dose (Table S3) with no detectable anti-N IgG in 44.4% (4/9) participants just prior to receiving their 2^nd^ vaccine dose.

### Anti-Spike IgG and neutralising antibody detection and quantification

The antibody positivity rate for all three S-protein based commercial assays (RBD IgG, Trimeric S IgG, and GenScript nAb) increased on average by 87.2% (range 73.8-94.3%) between baseline and the 2-week samples in the BNT162b2 group, reaching levels of >88% positivity (p<0.0001 for each assay) just prior to the second vaccine dose (Fig. 1). Similarly, large positivity increases between 23.8 and 26.7% were observed between the 2-week post-first dose and pre-2^nd^ dose time points in the mRNA-1273 group with overall increases of >50% and >70% positivity at the two time points respectively, p<0.05 for all comparisons (Table 2). For both time points and with both vaccines the number of participants who were positive for nAbs was significantly lower than those who were positive for IgG binding antibodies by a factor of 10% to 20% (Fig. 1).

**Table 2.** Quantitative titers of testing using commercial assays.

| Group | Blood sample timepoint | Quantitative titers<sup>a</sup> |  |  |
| --- | --- | --- | --- | --- |
|  |  | RBD IgG Titer (AU/mL) | Trimeric S IgG Titer (AU/mL)<sup>c</sup> | nAb Titer (ng/mL) |
| BNT162b2 | Baseline | 0<sup>b</sup><br>(0-1296.6) | 1.85<sup>b</sup><br>(1.85-176) | 0<sup>b</sup><br>(0-1634.7) |
|  | 2-weeks | 653.3<br>(300.4-1497.5) | 166<br>(82.6-272) | 355.7<br>(161.6-567.5) |
|  | Pre-2<sup>nd</sup> dose | 1180<br>(687.-2135.2) | 195<br>(138-318.5) | 628.6<br>(397-954.6) |
| mRNA-1273 | 2-weeks | 157.7<br>(22.6-4083.5) | 27.6<br>(2.1-235.5) | 85.6<br>(0-462.4) |
|  | Pre-2<sup>nd</sup> dose | 557.3<br>(167.9-8210.6) | 112<br>(34.5-764) | 381.1<br>(11.9-8802.8) |
| P-values |  |  |  |  |
| BNT162b2<sup>d</sup> | Baseline vs. 2 weeks | <0.0001 | <0.0001 | <0.0001 |
|  | 2 weeks vs. Pre-2<sup>nd</sup> dose | <0.0001 | <0.05 | <0.0001 |
| mRNA-1273 | 2 weeks vs Pre-2<sup>nd</sup> dose | 0.06 | <0.01 | <0.05 |

<sup>a</sup>Expressed as median value (IQR), except for BNT162b2 baseline samples.<sup>b</sup>Expressed as median value (range) due to low numbers of positive samples.<sup>c</sup>According to product insert, titers <13 AU/mL (negative) and ≥13 AU/mL (positive).
<sup>d</sup>All p-values comparing baseline to pre-2nd dose for the BNT162b2 vaccine <0.0001, except for Nucleocapsid IgG where p-value=1.0; NS= Not Significant.

Quantifiable titers of antibodies detected by each assay were detectable at all time points and increased significantly over time in both vaccine groups (Table 2 and Fig. 2). In the BNT162b2 group, titers increased by 1.8, 1.2, and 1.8-fold for the RBD IgG, Trimeric S IgG, and GenScript nAb assays respectively, from the 2-week blood sampling to the samples drawn just prior to the participants’ 2^nd^ dose (with a median number of 11 days between blood draws). Higher-fold titer increases (3.5, 4.1, and 4.5-fold respectively) were seen in the mRNA-1273 participants between blood sampling at similar time points (p=0.06 for RBD IgG and p<0.05 for Trimeric S IgG and nAb assays) (median seven days between blood draws) (Table 2 and Fig. 1). Titer values for a given group and time point were distributed across varying IQRs despite being all classified as positive (Fig. S1).

**Figure 2.**
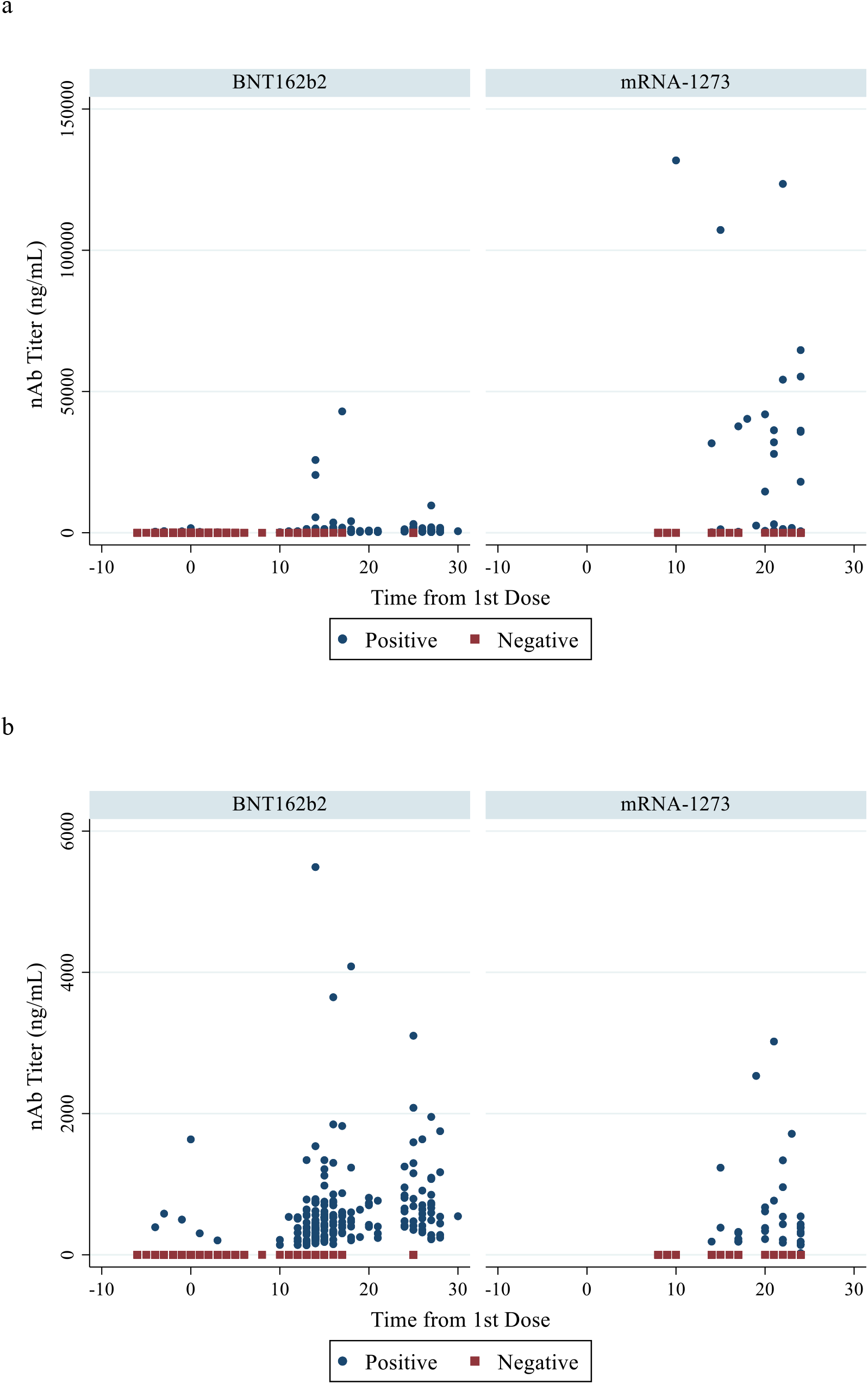
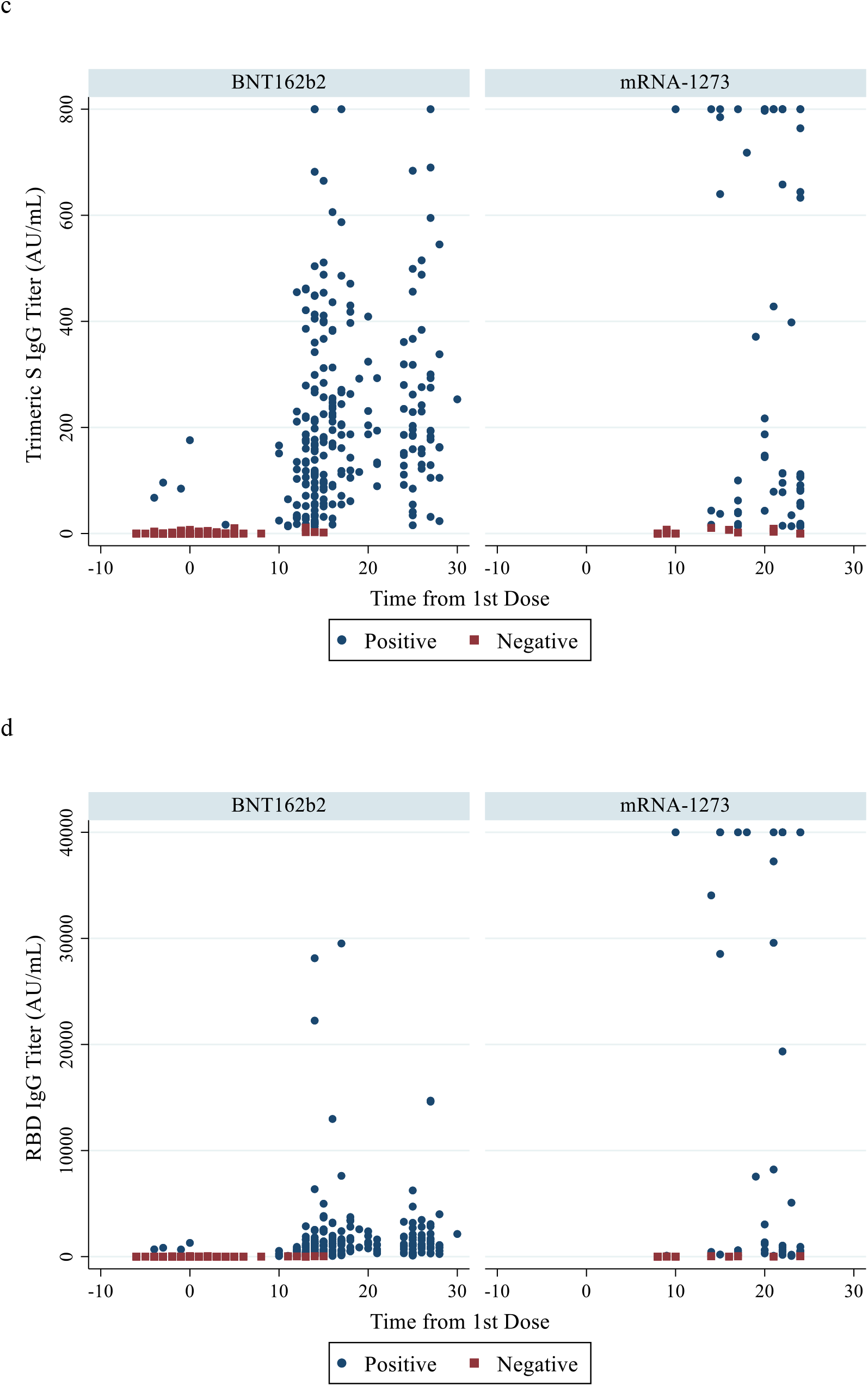
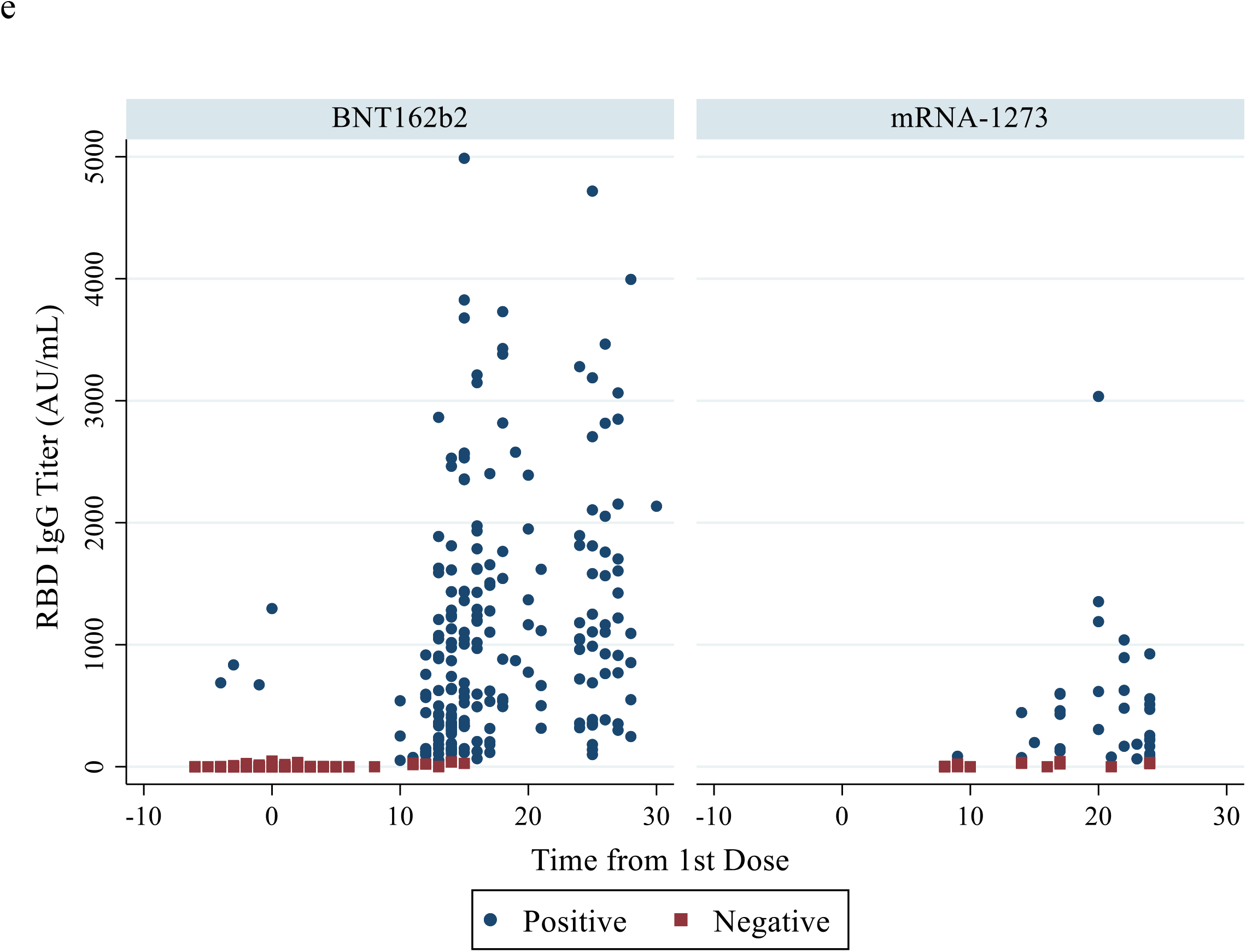
Quantitative testing results from time of first vaccine dose for each vaccine group using the nAb assay (panels a and b); Trimeric S IgG assay (panel c); and RBD IgG assay (panels d and e). Panel (b) presents panel (a) results with y-axis limited to maximum titer of 6000 ng/mL. Panel (e) presents panel (d) results with a shortened y-axis (max titer 5000 AU/mL).

### Post vaccine testing in special populations

Quantitative titres in the mRNA-1273 group were found to be significantly higher among participants who were COVID-19 recovered compared to those with no history of laboratory-confirmed COVID-19 infection (Table S4; *p*<0.001 for all assays at both time points). This was observed for the two commercial anti-S IgG assays as well as the nAb assay used. This comparison was made only in the mRNA-1273 group given all COVID-19 recovered participants received their first dose of vaccine >14 days from the date of their COVID-19 diagnosis.

Our sample included a small number of participants (n=5; four from the BNT162b2 group) on biologic or immunosuppressant therapy (Table S5). Four had no evidence of previous COVID-19 infection at baseline. In the group of four (who all received the BNT162b2 vaccine), quantitative anti-S IgG/nAb were detected at 2-weeks (4/4) whereas only two samples gave detectable levels of neutralising antibodies and the other two were close to baseline. The absolute values of quantitative titres were comparatively larger for the one participant who was COVID-19 recovered (mRNA-1273 group).

## Discussion

In this study, we demonstrate that vaccine-induced antibodies stimulated by FDA approved SARS-CoV-2 mRNA vaccines (BNT162b2 and mRNA-1273) can be detected by commercial high-throughput assays currently used in clinical laboratories. As predicted, detectable antibody was restricted to assays identifying IgG directed against the viral S-protein, and not against the nucleocapsid protein. Anti-spike vaccine-induced antibodies were detectable as early as ten days following the first dose of an mRNA vaccine. Titres increased significantly over the three-week period from baseline sampling to pre-second dose sampling. Vaccine-induced nAbs targeting the SARS-CoV-2 spike protein were successfully detected, using a surrogate virus neutralization test (sVNT) kit without the use of cell lines and viral culture required for plaque reduction neutralization assays (PRNT) supporting protective immunity [7, 8, 10, 22]. However, there were significantly less participants with detectable nAbs post-vaccination as those with a binding antibody IgG response.

To our knowledge, detection of vaccine-induced antibodies using commercial high-throughput SARS-CoV-2 anti-S IgG assays has been limited to a partnership between Roche® and Moderna® demonstrating detection of mRNA-1273 vaccine-induced antibodies using the Elecsys anti-SARS-CoV-2 S test [15] two other studies [23, 24]. Our current study adds to the literature by confirming the detection of these IgG binding and also functional nAbs using commercial kits from other companies likely to be adopted by large volume clinical laboratories.

Overall, our data demonstrate significant increases in the median titre of anti-S IgG by all assays evaluated in both mRNA vaccine groups, except for the RBD IgG among the mRNA-1273 group which did not measure levels from baseline but only from two weeks post vaccination. Given the lack of an international standard, antibody titres can only be directly compared between successive samples measured on the same assay run [25]. However, when samples from participants in the same vaccine group on the same assay were compared, wide IQRs of titre values were observed (Table 2; Fig. S1). The observed variation is likely multifactorial, and dependant on history of previous SARS-CoV-2 infection, use of immunosuppressive medications, advanced age, and frailty [26]. There is limited data to conclude whether the differing titres in participants in the same group (classified qualitatively as positive), assay, and time-point of sampling, translates to clinical differences in response to infection. For other vaccines, such as hepatitis B and varicella, an initial antibody response following first vaccine dose generally translates into protection from subsequent infection [27, 28]. However, protection is derived from both humoral and cell-mediated responses, of which only humoral is detected by the commercial high throughput EIAs [27, 28].

For other viral infections including SARS-CoV-2, there is significant evidence that nAbs are correlated to protection and immunity [7, 11]. Animal challenge studies have provided conclusive evidence for nAb protection to SARS-CoV-2 [22]. nAbs have also been successfully tested in humans to treat the disease [29]. When comparing the prevalence of neutralising and IgG antibodies from SARS-CoV-2 infected and recovered individuals, the prevalence of nAbs was comparable to IgG binding antibodies and to gold standard PRNT [30]. However, we found a significantly lower fraction of participants elicited protective nAbs compared to those with IgG antibodies. A potential explanation is RBD IgG and trimeric S IgG assays measure total binding antibodies, but are unable to differentiate between nAbs and non-nAbs that also bind to the S-protein [31]. Several studies comparing surrogate nAb assays to traditional methods (PRNT) have found them to be effective proxies for detection and quantification of immunodominant nAbs seen in SARS-CoV-2 infection, specifically RBD-targeting nAbs [21, 31] [32]. Encouragingly, at the final study blood draw (pre-second vaccination dose), nAbs were detectable in 73-89% of samples tested from both vaccine groups. Although the prevalence of nAbs remained significantly lower than total binding IgG’s, the data support that the majority of individuals would have at least some protection from subsequent infection prior to the second dose, particularly as detection of nAbs has been previously correlated with vaccine efficacy [33, 34]. However, these data underline a cautionary note in that up to 27% of vaccinated individuals did not have protective nAbs prior to the second dose and should remain vigilant in exercising protective public health measures to limit viral spread.

In a post-COVID-19-vaccination era, it may not be necessary to differentiate between vaccine induced antibody responses and natural infection. Although we found, at baseline, a number of COVID-19 recovered participants had detectable anti-N IgG (which would confirm previous infection), 30% were undetectable by the pre-second dose blood draw (Table S2 and Table S3). This may be due to a number of factors including, the severity of the initial SARS-CoV-2 infection [35], the specific anti-N IgG assay used [36], the natural decline of N antibodies overtime [37], or possible immune modulation from the vaccine, where the abundance of antibodies produced switched from anti-N following natural infection to anti-S antibodies following vaccination.

To effectively compare antibody titres (across vaccine groups and assays), a quantitative reference standard and a robust correlation with nAbs is needed for commercial assays to accurately assess the effectiveness of vaccine responses. It will be important for clinical laboratories to identify that vaccine induced antibodies can be detected for each vaccine, and work with public health partners to understand the best use for serological testing. Current guidelines [38, 39] do not recommend clinical serology testing pre or post-vaccination, however these recommendations may change over time with more study. A further understanding of the population-based efficacy and longevity is required to structure recommendations for individualized post-vaccine serology testing in clinical decision making.

Although sample sizes were low, we observed in the mRNA-1273 group that vaccine-induced antibody responses titres of all three quantitative assays were significantly higher at all time points in participants who recovered from confirmed COVID-19 infection (Table S4). Similar findings have been reported from several other research groups where post-vaccination titres were 10-20 times higher in COVID-19 recovered individuals [40, 41]. Furthermore, the responses prior to the second dose of vaccine in COVID-19 recovered participants also exceeded the titres of those from the SARS-CoV-2 naïve participants by 10-fold. This has led to early recommendations by some governments to consider providing only a one-dose primary series in COVID-19 recovered individuals [42].

For the four participants on immune suppressive treatment who were not previously SARS-CoV-2 infected, a marked difference between nAb and IgG response to vaccination was observed. Although the participants in this group all produced measurable IgG’s two weeks after receiving the first vaccine dose, only two gave measurable nAb levels and the remaining two were just above the detection limit. These data underline the importance of assuring immune protection to SARS-CoV-2 in this highly susceptible subpopulation of immunocompromised individuals.

The principal limitation in our findings is that they only demonstrate the production of vaccine-induced antibodies in response to mRNA COVID-19 vaccines and their quantification. Correlations of specific titre levels to protection or vaccine efficacy cannot be made as the study is not designed or powered for this purpose. As the BNT162b2 and mRNA-1273 vaccines are different products, it is difficult to comment on the difference in median titres seen for the same time points on the same assay (which was observed in general to be lower in the mRNA-1273 group, despite a higher proportion of COVID-19 recovered individuals). This may reflect differing median ages of the two groups or alternatively be reflective of responses to varying vaccine products which is an artifact that could be corrected with an international standard for COVID-19 serology.

## Conclusions

We have shown that currently available commercial assays for detection of anti-spike IgG binding antibodies were successfully able to detect vaccine induced antibodies in the vast majority of vaccinated participants. In general, anti-spike IgG binding antibody titers increased overtime, while anti-nucleocapsid antibodies declined. There was a marked difference in the proportion of participants eliciting protective nAbs for both vaccines and at all time points after the first dose underlining the difference between a general binding IgG and a functional neutralising response correlating to protection. Just prior to the second dose of vaccination, most samples had detectable neutralising antibodies. However, a significant subpopulation did not produce nAbs which underlines the importance of the continued practice of SARS-CoV-2 infection preventive measures after the first vaccine dose at least for those individuals with no detectable levels of nAbs.

## Supporting information

STROBE checklist

## Data Availability

The datasets used and/or analysed during the current study are not publicly available.

## Declarations

### Research ethics approval

This study was approved by the Human Research Ethics Board (HREB) at the University of Alberta (reference number Pro00106810).

### Consent to publication

Not applicable.

### Conflicts of interest/Competing interests

Sean Taylor is an employee of GenScript USA. He assisted in providing interpretation of the GenScript assay used in this study. He had no role in the design of the study or the formal data analysis, but did provide suggestions for the final draft which were incorporated after review by the other authors. None of the other authors had conflicts to declare with respect to this manuscript.

### Funding

This work did not receive any specific grant from funding agencies in the public, commercial, or not-for-profit sectors, however testing kits were provided in-kind by all manufactures.

### Availability of data and material

The datasets used and/or analysed during the current study are available from the corresponding author on reasonable request.

### Authors’ contributions

Jamil N Kanji – Conceptualization, methodology, validation, analysis, data curation, project administration, writing – original draft, writing – review and editing.

Ashley Bailey – validation, investigation, data curation, writing – review and editing

Jayne Fenton – investigation, data curation, writing – review and editing

Sean H Ling – investigation, data curation, project administration, writing – review and editing

Rafael Rivera – data curation, formal analysis, writing – review and editing

Sabrina Plitt – data curation, formal analysis, project administration, writing – review and editing

Wendy I Sligl – data curation, investigation, writing – review and editing, project administration

Sean Taylor – resources, writing – review and editing

LeeAnn Turnbull – investigation, writing – review and editing

Graham Tipples – project administration, writing – review and editing

Carmen Charlton – Conceptualization, methodology, validation, resources, writing – original draft, writing – review and editing, supervision, project administration

## Acknowledgements

We would like to thank Abbott Laboratories, DiaSorin and GenScript for providing us with complimentary serology testing kits.

## ICMJE Criteria

All authors attest they meet the ICMJE criteria for authorship.

## Notes

### Competing Interest Statement

Dr. Sean Taylor is an employee of GenScript USA. He assisted in providing interpretation of the GenScript assay used in this study. He had no role in the design of the study or the formal data analysis, but did provide suggestions for the final draft which were incorporated after review by the other authors. None of the other authors had conflicts to declare with respect to this manuscript.

## References

[1] Van Caeseele P, Bailey D, Forgie SE, Dingle TC, Krajden M (2020) SARS-CoV-2 (COVID-19) serology: implications for clinical practice, laboratory medicine and public health. Cmaj 192 (34):E973–e979

[2] Rostami A, Sepidarkish M, Leeflang MMG, Riahi SM, Nourollahpour Shiadeh M, Esfandyari S, Mokdad AH, Hotez PJ, Gasser RB (2020) SARS-CoV-2 seroprevalence worldwide: a systematic review and meta-analysis. Clin Microbiol Infect

[3] Sahin U, Muik A, Derhovanessian E, Vogler I, Kranz LM, Vormehr M, Baum A, Pascal K, Quandt J, Maurus D, Brachtendorf S, Lörks V, Sikorski J, Hilker R, Becker D, Eller A-K, Grützner J, Boesler C, Rosenbaum C, Kühnle M-C, Luxemburger U, Kemmer-Brück A, Langer D, Bexon M, Bolte S, Karikó K, Palanche T, Fischer B, Schultz A, Shi P-Y, Fontes-Garfias C, Perez JL, Swanson KA, Loschko J, Scully IL, Cutler M, Kalina W, Kyratsous CA, Cooper D, Dormitzer PR, Jansen KU, Türeci Ö (2020) COVID-19 vaccine BNT162b1 elicits human antibody and TH1 T cell responses. Nature 586 (7830):594–599

[4] Barrett JR, Belij-Rammerstorfer S, Dold C, Ewer KJ, Folegatti PM, Gilbride C, Halkerston R, Hill J, Jenkin D, Stockdale L, Verheul MK, Aley PK, Angus B, Bellamy D, Berrie E, Bibi S, Bittaye M, Carroll MW, Cavell B, Clutterbuck EA, Edwards N, Flaxman A, Fuskova M, Gorringe A, Hallis B, Kerridge S, Lawrie AM, Linder A, Liu X, Madhavan M, Makinson R, Mellors J, Minassian A, Moore M, Mujadidi Y, Plested E, Poulton I, Ramasamy MN, Robinson H, Rollier CS, Song R, Snape MD, Tarrant R, Taylor S, Thomas KM, Voysey M, Watson MEE, Wright D, Douglas AD, Green CM, Hill AVS, Lambe T, Gilbert S, Pollard AJ, Aboagye J, Alderson J, Ali A, Allen E, Allen L, Anslow R, Arancibia-Cárcamo CV, Arbe-Barnes EH, Baker M, Baker P, Baker N, Baleanu I, Barnes E, Bates L, Batten A, Beadon K, Beckley R, Beveridge A, Bewley KR, Bijker EM, Blackwell L, Blundell CL, Bolam E, Boland E, Borthwick N, Boyd A, Brenner T, Brown P, Brown-O’Sullivan C, Brunt E, Burbage J, Buttigieg KR, Byard N, Cabrera Puig I, Camara S, Cao M, Cappuccini F, Carr M, Carroll MW, Chadwick J, Chelysheva I, Cho J-S, Cifuentes L, Clark E, Colin-Jones R, Conlon CP, Coombes NS, Cooper R, Crocker WEM, Cunningham CJ, Damratoski BE, Datoo MS, Datta C, Davies H, Demissie T, Di Maso C, DiTirro D, Dong T, Donnellan FR, Douglas N, Downing C, Drake J, Drake-Brockman R, Drury RE, Dunachie SJ, Muhanna OE, Elias SC, Elmore MJ, Emary KRW, English MR, Felle S, Feng S, Da Silva CF, Field S, Fisher R, Ford KJ, Fowler J, Francis E, Frater J, Furze J, Galian-Rubio P, Garlant H, Godwin K, Gorini G, Gracie L, Gupta G, Hamilton E, Hamlyn J, Hanumunthadu B, Harris SA, Harrison D, Hart TC, Hawkins S, Henry JA, Hodges G, Hodgson SHC, Hou MM, Howe E, Howell N, Huang B, Humphries H, Iveson P, Jackson S, Jackson F, Jauregui S, Jeffery K, Jones E, Jones K, Kailath R, Keen J, Kelly S, Kelly D, Kelly E, Kerr D, Khan L, Khozoee B, Killen A, Kinch J, King TB, King L, Kingham-Page L, Klenerman P, Knight JC, Knott D, Koleva S, Larkworthy CW, Larwood JPJ, Lees EA, Lelliott A, Leung S, Li Y, Lias AM, Lipworth S, Liu S, Loew L, Lopez Ramon R, Mallett G, Mansatta K, Marchevsky NG, Marinou S, Marlow E, Marshall JL, Matthews P, McEwan J, McGlashan J, McInroy L, Meddaugh G, Mentzer AJ, Mirtorabi N, Morey E, Morgans R, Morris SJ, Morrison H, Morshead G, Morter R, Moya N, Mukhopadhyay E, Muller J, Munro C, Murphy S, Mweu P, Noé A, Nugent FL, Nuthall E, O’Brien K, O’Connor D, O’Donnell D, Oguti B, Olchawski V, Oliveria C, O’Reilly PJ, Osborne P, Owino N, Parker K, Parracho H, Patrick-Smith M, Peng Y, Penn E, Peralta Alvarez MP, Perring J, Petropoulos C, Pfafferott K, Pipini D, Phillips D, Proud P, Provstgaard-Morys S, Pulido D, Radia K, Rajapaksa D, Ramos Lopez F, Ratcliffe H, Rawlinson T, Pabon ER, Rhead S, Ritchie AJ, Roberts H, Roche S, Rudiansyah I, Salvador S, Sanders H, Sanders K, Satti I, Schmid A, Schofield E, Screaton G, Sedik C, Shaik I, Sharpe HR, Shea A, Silk S, Silva-Reyes L, Skelly DT, Smith CC, Smith DJ, Spencer AJ, Stafford E, Szigeti A, Tahiri-Alaoui A, Tanner R, Taylor IJ, Taylor K, te Water Naude R, Themistocleous Y, Themistocleous A, Thomas M, Thomas TM, Thompson A, Tinh L, Tomic A, Tonks S, Towner J, Tran N, Tree JA, Truby A, Turner C, Turner N, Ulaszewska M, Varughese R, Vichos I, Walker L, Wand M, White C, White R, Williams P, Worth AT, the Oxford CVTG (2020) Phase 1/2 trial of SARS-CoV-2 vaccine ChAdOx1 nCoV-19 with a booster dose induces multifunctional antibody responses. Nature Medicine

[5] Jackson LA, Anderson EJ, Rouphael NG, Roberts PC, Makhene M, Coler RN, McCullough MP, Chappell JD, Denison MR, Stevens LJ, Pruijssers AJ, McDermott A, Flach B, Doria-Rose NA, Corbett KS, Morabito KM, O’Dell S, Schmidt SD, Swanson PA, 2nd, Padilla M, Mascola JR, Neuzil KM, Bennett H, Sun W, Peters E, Makowski M, Albert J, Cross K, Buchanan W, Pikaart-Tautges R, Ledgerwood JE, Graham BS, Beigel JH (2020) An mRNA Vaccine against SARS-CoV-2 -Preliminary Report. N Engl J Med 383 (20):1920–1931

[6] Centers for Disease Control and Prevention Chickenpox (Varicella): Interpreting laboratory tests, https://www.cdc.gov/chickenpox/lab-testing/lab-tests.html. Cited January 1, 2021

[7] Garcia-Beltran WF, Lam EC, Astudillo MG, Yang D, Miller TE, Feldman J, Hauser BM, Caradonna TM, Clayton KL, Nitido AD, Murali MR, Alter G, Charles RC, Dighe A, Branda JA, Lennerz JK, Lingwood D, Schmidt AG, Iafrate AJ, Balazs AB (2021) COVID-19-neutralizing antibodies predict disease severity and survival. Cell 184 (2):476-488.e411

[8] Addetia A, Crawford KH, Dingens A, Zhu H, Roychoudhury P, Huang M-L, Jerome KR, Bloom JD, Greninger AL (2020) Neutralizing antibodies correlate with protection from SARS-CoV-2 in humans during a fishery vessel outbreak with high attack rate. Journal of Clinical Microbiology

[9] Hassan AO, Case JB, Winkler ES, Thackray LB, Kafai NM, Bailey AL, McCune BT, Fox JM, Chen RE, Alsoussi WB (2020) A SARS-CoV-2 infection model in mice demonstrates protection by neutralizing antibodies. Cell 182 (3):744-753. e744

[10] Yu J, Tostanoski LH, Peter L, Mercado NB, McMahan K, Mahrokhian SH, Nkolola JP, Liu J, Li Z, Chandrashekar A, Martinez DR, Loos C, Atyeo C, Fischinger S, Burke JS, Slein MD, Chen Y, Zuiani A, Lelis FJN, Travers M, Habibi S, Pessaint L, Van Ry A, Blade K, Brown R, Cook A, Finneyfrock B, Dodson A, Teow E, Velasco J, Zahn R, Wegmann F, Bondzie EA, Dagotto G, Gebre MS, He X, Jacob-Dolan C, Kirilova M, Kordana N, Lin Z, Maxfield LF, Nampanya F, Nityanandam R, Ventura JD, Wan H, Cai Y, Chen B, Schmidt AG, Wesemann DR, Baric RS, Alter G, Andersen H, Lewis MG, Barouch DH (2020) DNA vaccine protection against SARS-CoV-2 in rhesus macaques. Science (New York, NY) 369 (6505):806–811

[11] Plotkin SA (2010) Correlates of protection induced by vaccination. Clin Vaccine Immunol 17 (7):1055–1065

[12] Baden LR, El Sahly HM, Essink B, Kotloff K, Frey S, Novak R, Diemert D, Spector SA, Rouphael N, Creech CB, McGettigan J, Khetan S, Segall N, Solis J, Brosz A, Fierro C, Schwartz H, Neuzil K, Corey L, Gilbert P, Janes H, Follmann D, Marovich M, Mascola J, Polakowski L, Ledgerwood J, Graham BS, Bennett H, Pajon R, Knightly C, Leav B, Deng W, Zhou H, Han S, Ivarsson M, Miller J, Zaks T (2020) Efficacy and Safety of the mRNA-1273 SARS-CoV-2 Vaccine. New England Journal of Medicine

[13] Polack FP, Thomas SJ, Kitchin N, Absalon J, Gurtman A, Lockhart S, Perez JL, Pérez Marc G, Moreira ED, Zerbini C, Bailey R, Swanson KA, Roychoudhury S, Koury K, Li P, Kalina WV, Cooper D, Frenck RW, Hammitt LL, Türeci Ö, Nell H, Schaefer A, Ünal S, Tresnan DB, Mather S, Dormitzer PR, Şahin U, Jansen KU, Gruber WC (2020) Safety and Efficacy of the BNT162b2 mRNA Covid-19 Vaccine. New England Journal of Medicine 383 (27):2603–2615

[14] Sahin U, Muik A, Vogler I, Derhovanessian E, Kranz LM, Vormehr M, Quandt J, Bidmon N, Ulges A, Baum A, Pascal K, Maurus D, Brachtendorf S, Lörks V, Sikorski J, Koch P, Hilker R, Becker D, Eller A-K, Grützner J, Tonigold M, Boesler C, Rosenbaum C, Heesen L, Kühnle M-C, Poran A, Dong JZ, Luxemburger U, Kemmer-Brück A, Langer D, Bexon M, Bolte S, Palanche T, Schultz A, Baumann S, Mahiny AJ, Boros G, Reinholz J, Szabó GT, Karikó K, Shi P-Y, Fontes-Garfias C, Perez JL, Cutler M, Cooper D, Kyratsous CA, Dormitzer PR, Jansen KU, Türeci Ö (2020) BNT162b2 induces SARS-CoV-2-neutralising antibodies and T cells in humans. medRxiv:2020.2012.2009.20245175

[15] Roche Roche partners with Moderna to include SARS-CoV-2 antibody test in ongoing COVID-19 vaccine trials, https://www.roche.com/media/releases/med-cor-2020-12-09.htm. Cited January 17, 2021

[16] Widge AT, Rouphael NG, Jackson LA, Anderson EJ, Roberts PC, Makhene M, Chappell JD, Denison MR, Stevens LJ, Pruijssers AJ, McDermott AB, Flach B, Lin BC, Doria-Rose NA, O’Dell S, Schmidt SD, Neuzil KM, Bennett H, Leav B, Makowski M, Albert J, Cross K, Edara V-V, Floyd K, Suthar MS, Buchanan W, Luke CJ, Ledgerwood JE, Mascola JR, Graham BS, Beigel JH (2020) Durability of Responses after SARS-CoV-2 mRNA-1273 Vaccination. New England Journal of Medicine 384 (1):80–82

[17] Government of Alberta COVID-19 vaccine distribution, https://www.alberta.ca/covid19-vaccine.aspx. Cited February 4, 2021

[18] Abbott SARS-CoV-2 Immunoassays, https://www.corelaboratory.abbott/int/en/offerings/segments/infectious-disease/sars-cov-2. Cited February 21, 2021

[19] DiaSorin Liaison(R) SARS-CoV-2 TrimericS IgG Aassay, https://www.diasorin.com/sites/default/files/allegati_prodotti/liaisonr_sars-cov-2_trimerics_igg_assay_m0870004408.pdf. Cited February 21, 2021

[20] GenScript SARS-CoV-2 surrogate virus neutralization test (sVNT) kit, https://www.genscript.com/covid-19-detection-svnt.html. Cited February 21, 2021

[21] Taylor SC, Hurst B, Charlton CL, Bailey A, Kanji JN, McCarthy MK, Morrison TE, Huey L, Annen K, DomBourian MG, Knight V (2021) A New SARS CoV-2 Dual Purpose Serology Test: Highly Accurate Infection Tracing and Neutralizing Antibody Response Detection. J Clin Microbiol

[22] Rogers TF, Zhao F, Huang D, Beutler N, Burns A, He WT, Limbo O, Smith C, Song G, Woehl J, Yang L, Abbott RK, Callaghan S, Garcia E, Hurtado J, Parren M, Peng L, Ramirez S, Ricketts J, Ricciardi MJ, Rawlings SA, Wu NC, Yuan M, Smith DM, Nemazee D, Teijaro JR, Voss JE, Wilson IA, Andrabi R, Briney B, Landais E, Sok D, Jardine JG, Burton DR (2020) Isolation of potent SARS-CoV-2 neutralizing antibodies and protection from disease in a small animal model. Science 369 (6506):956–963

[23] Prendecki M, Clarke C, Brown J, Cox A, Gleeson S, Guckian M, Randell P, Pria AD, Lightstone L, Xu X-N, Barclay W, McAdoo SP, Kelleher P, Willicombe M Effect of previous SARS-CoV-2 infection on humoral and T-cell responses to single-dose BNT162b2 vaccine. The Lancet

[24] Manisty C, Otter AD, Treibel TA, McKnight Á, Altmann DM, Brooks T, Noursadeghi M, Boyton RJ, Semper A, Moon JC (2021) Antibody response to first BNT162b2 dose in previously SARS-CoV-2-infected individuals. Lancet

[25] Kempster SL, Almond N, Dimech W, Grangeot-Keros L, Huzly D, Icenogle J, El Mubarak HS, Mulders MN, Nübling CM (2020) WHO international standard for anti-rubella: learning from its application. Lancet Infect Dis 20 (1):e17–e19

[26] Andrew MK, McElhaney JE (2020) Age and frailty in COVID-19 vaccine development. The Lancet 396 (10267):1942–1944

[27] Spradling PR, Xing J, Williams R, Masunu-Faleafaga Y, Dulski T, Mahamud A, Drobeniuc J, Teshale EH (2013) Immunity to hepatitis B virus (HBV) infection two decades after implementation of universal infant HBV vaccination: association of detectable residual antibodies and response to a single HBV challenge dose. Clin Vaccine Immunol 20 (4):559–561

[28] Greub G, Zysset F, Genton B, Spertini F, Frei PC (2001) Absence of anti-hepatitis B surface antibody after vaccination does not necessarily mean absence of immune response. Med Microbiol Immunol 189 (3):165–168

[29] Weinreich DM, Sivapalasingam S, Norton T, Ali S, Gao H, Bhore R, Musser BJ, Soo Y, Rofail D, Im J (2021) REGN-COV2, a neutralizing antibody cocktail, in outpatients with Covid-19. New England Journal of Medicine 384 (3):238–251

[30] Perera RAPM, Ko R, Tsang OTY, Hui DSC, Kwan MYM, Brackman CJ, To EMW, Yen H-l, Leung K, Cheng SMS, Chan KH, Chan KCK, Li K-C, Saif L, Barrs VR, Wu JT, Sit THC, Poon LLM, Peiris M (2020) Evaluation of a SARS-CoV-2 surrogate virus neutralization test for detection of antibody in human, canine, cat and hamster sera. Journal of Clinical Microbiology:JCM.02504-02520

[31] Tan CW, Chia WN, Qin X, Liu P, Chen MIC, Tiu C, Hu Z, Chen VC-W, Young BE, Sia WR, Tan Y-J, Foo R, Yi Y, Lye DC, Anderson DE, Wang L-F (2020) A SARS-CoV-2 surrogate virus neutralization test based on antibody-mediated blockage of ACE2–spike protein–protein interaction. Nature Biotechnology 38 (9):1073–1078

[32] Premkumar L, Segovia-Chumbez B, Jadi R, Martinez DR, Raut R, Markmann AJ, Cornaby C, Bartelt L, Weiss S, Park Y, Edwards CE, Weimer E, Scherer EM, Rouphael N, Edupuganti S, Weiskopf D, Tse LV, Hou YJ, Margolis D, Sette A, Collins MH, Schmitz J, Baric RS, de Silva AM (2020) The receptor-binding domain of the viral spike protein is an immunodominant and highly specific target of antibodies in SARS-CoV-2 patients. Science Immunology 5 (48):eabc8413

[33] Amit S, Regev-Yochay G, Afek A, Kreiss Y, Leshem E Early rate reductions of SARS-CoV-2 infection and COVID-19 in BNT162b2 vaccine recipients. The Lancet

[34] Pawlowski C, Lenehan P, Puranik A, Agarwal V, Venkatakrishnan A, Niesen MJ, O Horo JC, Badley AD, Halamka J, Soundararajan V (2021) FDA-authorized COVID-19 vaccines are effective per real-world evidence synthesized across a multi-state health system. medRxiv:2021.2002.2015.21251623

[35] Bruni M, Cecatiello V, Diaz-Basabe A, Lattanzi G, Mileti E, Monzani S, Pirovano L, Rizzelli F, Visintin C, Bonizzi G, Giani M, Lavitrano M, Faravelli S, Forneris F, Caprioli F, Pelicci PG, Natoli G, Pasqualato S, Mapelli M, Facciotti F (2020) Persistence of Anti-SARS-CoV-2 Antibodies in Non-Hospitalized COVID-19 Convalescent Health Care Workers. J Clin Med 9 (10)

[36] Muecksch F, Wise H, Batchelor B, Squires M, Semple E, Richardson C, McGuire J, Clearly S, Furrie E, Greig N, Hay G, Templeton K, Lorenzi JCC, Hatziioannou T, Jenks S, Bieniasz PD (2021) Longitudinal Serological Analysis and Neutralizing Antibody Levels in Coronavirus Disease 2019 Convalescent Patients. J Infect Dis 223 (3):389–398

[37] Mai HK, Trieu NB, Long TH, Thanh HT, Luong ND, Huy LX, Nguyet LA, Man DNH, Anderson DE, Thanh TT, Chau NVV, Thwaites G, Wang LF, Van Tan L, Hung DT (2021) Long-Term Humoral Immune Response in Persons with Asymptomatic or Mild SARS-CoV-2 Infection, Vietnam. Emerg Infect Dis 27 (2):663–666

[38] Government of Canada National Advisory Committee on Immunization (NACI): Recommendations on the use of COVID-19 vaccines, https://www.canada.ca/en/public-health/services/immunization/national-advisory-committee-on-immunization-naci/recommendations-use-covid-19-vaccines.html. Cited March 5, 2021

[39] Centers for Disease Control and Prevention Vaccine Recommendations and Guidelines of the Advisory Committee on Immunization Practices (ACIP): COVID-19 ACIP Vaccine Recommendations, https://www.cdc.gov/vaccines/hcp/acip-recs/vacc-specific/covid-19.html. Cited March 5, 2021

[40] Krammer F, Srivastava K, Simon V (2021) Robust spike antibody responses and increased reactogenicity in seropositive individuals after a single dose of SARS-CoV-2 mRNA vaccine. medRxiv:2021.2001.2029.21250653

[41] Saadat S, Tehrani ZR, Logue J, Newman M, Frieman MB, Harris AD, Sajadi MM (2021) Binding and Neutralization Antibody Titers After a Single Vaccine Dose in Health Care Workers Previously Infected With SARS-CoV-2. JAMA

[42] British Broadcasting Corporation (BBC) Covid: France says just one jab needed for previously infected, https://www.bbc.com/news/world-europe-56048444. Cited February 25, 2021

