## Supplementary tables and figure. for "Detection of SARS-CoV-2 antibodies formed in response to the BNT162b2 and mRNA-1237 mRNA vaccine by commercial antibody tests"

### Supplementary information – Table of contents

**Table S1.** Commercial products used in this study.

**Table S2.** Anti-Nucleocapsid IgG detection in relation to history of confirmed laboratory diagnosis of SARS-CoV-2 infection in participants of BNT162b2 group (n = 6).

**Table S3.** Anti-Nucleocapsid IgG detection in relation to history of confirmed laboratory diagnosis of SARS-CoV-2 infection in participants of mRNA-1273 group (n = 16).

**Table S4.** Comparison of mean titers in mRNA-1273 group participants based on a history of previous COVID-19.

**Table S5**. Quantitative titers on each assay in patients on biologics/immunosuppressants.

**Fig. S1.** Box-whisker plots demonstrating the spread of only positive quantitative (titer) results for each vaccine group for the (a) RBD IgG assay; (b) Trimeric S IgG assay; and (c) nAb assay.

**Table S1.** Commercial products used in this study.

| Item | Description | Commercial affiliation |
|---|---|---|
| BNT162b2 vaccine | mRNA vaccine | Pfizer (New York, NY, USA) |
| mRNA-1273 vaccine | mRNA vaccine | Moderna Messenger Therapeutics (Cambridge, MA, USA) |
| Serology assay (Abbott Architect) | SARS-CoV-2 IgG targeting the nucleocapsid protein | Abbott (Chicago, IL, USA) |
| Serology assay (Abbott Architect) | SARS-CoV-2 IgG targeting the receptor binding domain (RBD) | Abbott (Chicago, IL, USA) |
| Serology assay (DiaSorin Liaison) | SARS-CoV-2 IgG targeting the S1/S2 regions of the spike protein | DiaSorin (Saluggia, Italy) |
| Serology assay (manual assay) | SARS-CoV-2 neutralizing antibody (nAb) preventing RBD and angiotensin-converting enzyme 2 (ACE-2) | GenScript (Piscataway, NJ, USA) |
| Electronic medical record (EMR) | Epic EMR | Epic (Verona, WI, USA) |
| Statistical software | Stata Version 16.1 | StataCorp (College Station, TX, USA) |

**Table S2.** Anti-Nucleocapsid IgG detection in relation to history of confirmed laboratory diagnosis of SARS-CoV-2 infection in participants of BNT162b2 group (n = 6).

| Case | Days between COVID-19 diagnosis and first vaccine dose[a] | Anti-N IgG result at baseline[b] | Anti-N IgG result 2-weeks post first dose of vaccine | Anti-N IgG result prior to second dose of vaccine |
|---|---|---|---|---|
| Case 1 | 27 | + | + | ND |
| Case 2 | 45 | + | + | + |
| Case 3 | 46 | + | - | ND |
| Case 4 | 118 | + | + | ND |
| Case 5 | -7 | - | ND | + |
| Case 6[c] | -34 | - | - | - |

Abbreviations: ND – not done; + positive; - negative
[a]A negative value refers to the diagnosis of COVID-19 being made after the first dose of vaccine. All SARS-CoV-2 diagnosis were confirmed using PCR testing from a nasopharyngeal or oropharyngeal swab collected due to symptoms.
[b]Qualitative results on the RBD IgG, Trimeric S IgG, and GenScript nAb assays were concordant with the Anti-N IgG result at baseline for all six cases.
[c]Confirmed COVID-19 positive nine days after second dose of BNT162b2 vaccine.

**Table S3.** Anti-Nucleocapsid IgG detection in relation to history of confirmed laboratory diagnosis of SARS-CoV-2 infection in participants of mRNA-1273 group (n = 16).

| Case | Days between COVID-19 diagnosis and first vaccine dose[a] | Anti-N IgG result 2-weeks post first vaccine dose[b] | Anti-N IgG result prior to 2nd dose of vaccine[b] |
|---|---|---|---|
| Case 1 | 18 | + | ND |
| Case 2 | 18 | ND | ND |
| Case 3 | 22 | ND | ND |
| Case 4 | 27 | + | + |
| Case 5 | 34 | ND | + |
| Case 6 | 35 | ND | ND |
| Case 7 | 41 | ND | - |
| Case 8 | 41 | ND | ND |
| Case 9 | 45 | + | + |
| Case 10 | 45 | ND | ND |
| Case 11 | 48 | ND | ND |
| Case 12 | 73 | + | + |
| Case 13 | 73 | + | - |
| Case 14 | 73 | - | - |
| Case 15 | 76 | + | - |
| Case 16 | 76 | ND | + |

Abbreviations: ND – not done due to shortage of assay reagents ; + positive; - negative
[a]All SARS-CoV-2 diagnosis were confirmed using PCR testing from a nasopharyngeal or oropharyngeal swab collected due to symptoms.
[b]All of these patients were found to be positive in one or more of either RBD IgG, Trimeric S IgG, or nAb titer.

**Table S4.** Comparison of median titers (IQRs) in mRNA-1273 group participants based on a history of previous COVID-19.

| Time point | Assay | COVID-19 Recovered | No prior history of COVID-19 | p-value |
|---|---|---|---|---|
| 2-weeks post first vaccine dose | Trimeric S-IgG (AU/mL) | 785 (640-800) (n = 7) | 14.4 (0-40.3) (n = 25) | 0.0002 |
| | RBD IgG (AU/mL) | 40,000 (28,534.6-40,000) (n = 7) | 74.7 (3.6-429.7) (n = 25) | <0.0001 |
| | nAb (ng/mL) | 37,672 (1,233.8-107,194.2) (n = 7) | 0 (0-189.6) (n = 25) | 0.0009 |
| Pre-2nd vaccine dose | Trimeric S-IgG (AU/mL) | 800 (651-800) (n = 16) | 58.2 (13.8-113) (n = 31) | <0.0001 |
| | RBD IgG (AU/mL) | 40000 (37257.3-40000) (n = 9) | 280.9 (88.1-909.5) (n = 28) | <0.0001 |
| | nAb (ng/mL) | 33,874.2 (2,179.4-48,052.8) (n = 16) | 185.4 (0-386.2) (n = 32) | <0.0001 |

**Table S5**. Quantitative titers on each assay in patients on biologics/immunosuppressants.

| Timing of blood sample / Assay | Participant 1 (BNT162b2) | Participant 2 (BNT162b2) | Participant 3 (BNT162b2) | Participant 4 (BNT162b2) | Participant 5[a] (mRNA-1273) |
|---|---|---|---|---|---|
| Baseline blood sample | | | | | |
| RBD IgG (AU/mL) | 6.8 | 0 | 0 | 0 | ND |
| Trimeric S IgG (AU/mL) | 1.9 | 1.9 | 1.9 | 1.9 | ND |
| nAb (ng/mL) | 0 | 0 | 0 | 0 | ND |
| 2-week blood sample | | | | | |
| RBD IgG (AU/mL) | 270.6 | 127.4 | 492.9 | 641.4 | 7,541.3 |
| Trimeric S IgG (AU/mL) | 56.3 | 17.4 | 119 | 139 | 371 |
| nAb (ng/mL) | 0 | 0 | 477 | 250.2 | 2,534.2 |
| Pre-2nd dose blood sample | | | | | |
| RBD IgG (AU/mL) | ND | 247.2 | ND | 1,092.8 | 8,210.6 |
| Trimeric S IgG (AU/mL) | ND | 23.4 | ND | 162 | 428 |
| nAb (ng/mL) | ND | 281.4 | ND | 540.2 | 3,020.4 |

Abbreviations: ND – not done.

[a]Participant 5 is COVID-19 recovered (received first dose of mRNA-1273 vaccine 73 days post date of COVID-19 diagnosis).

**Figure S1:**

a

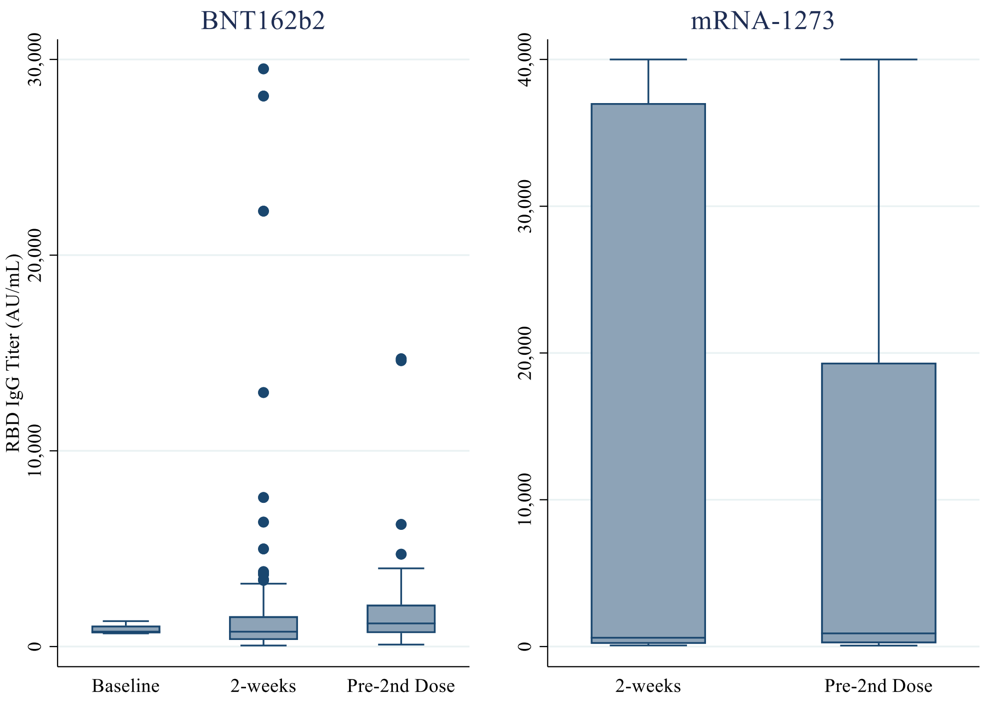

b

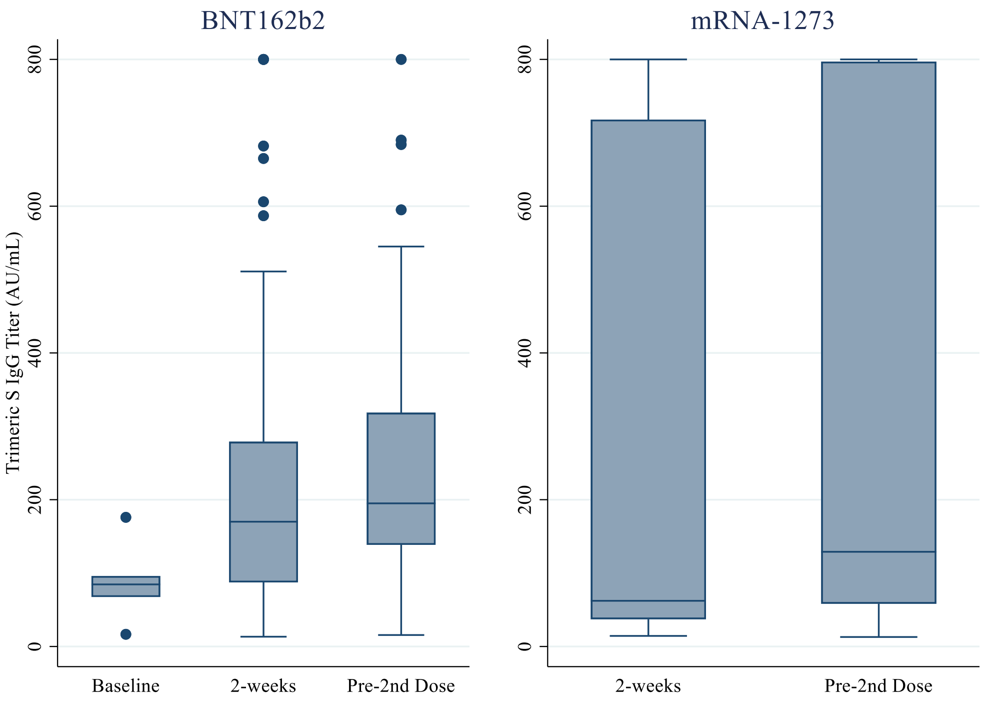

c

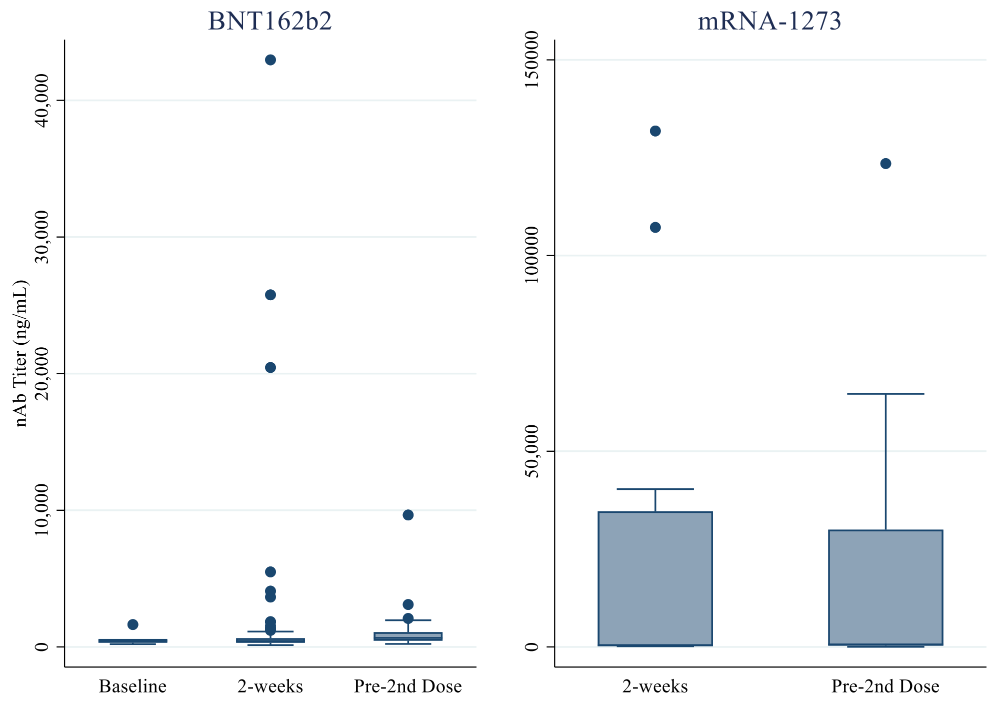

**Fig. S1.** Box-whisker plots demonstrating the spread of only positive quantitative (titer) results for each vaccine group for the (a) RBD IgG assay; (b) Trimeric S IgG assay; and (c) nAb assay.
